# Precise In Vivo Base-Editing Therapy for Alpha-1 Antitrypsin Deficiency

**DOI:** 10.64898/2026.11.25.26368393

**Authors:** Taihua Yang, Yanhong Jiang, Simon Alexander Krooss, Nora Drick, Jiahao Ge, Hongling Zhang, Fangfang Chen, Wensu Yu, Lifeng Yang, Xiaoying Ma, Qian Han, Julia Held, Heiner Wedemeyer, Ying Liu, Louisa Lilly Aurelia Ott, Rembert Koczulla, Sabina Janciauskiene, Marius Hoeper, Zi Jun Wang, Qinggong Yuan, Yuxuan Wu, Michael Ott, Qiang Xia

**Affiliations:** Department of Liver Surgery, Ren Ji Hospital, Shanghai Jiao Tong University School of Medicine, 1630 Dongfang Road, Shanghai, 200127, China; Shanghai Key Laboratory of Precision Gene Editing and Clinical Translation, Ren Ji Hospital, Shanghai Jiao Tong University School of Medicine, 1630 Dongfang Road, Shanghai, 200127, China; YolTech Therapeutics, 58 Tanzhu Road, Shanghai, 201109, China; Department of Gastroenterology, Hepatology, Infectious Diseases and Endocrinology, Hannover Medical School, Hannover, Germany; PRACTIS Clinician Scientist Program, Dean’s Office for Academic Career Development, Hannover Medical School, Hannover, Germany; Sino-German Gene and cell therapy center, Ren Ji Hospital, Shanghai, China; Department of Respiratory Medicine and Infectious Diseases, Hannover Medical School, Hannover, Germany; Member of the German Center for Lung Research (DZL), Biomedical Research in Endstage and Obstructive Lung Disease Hanover (BREATH), Hannover, Germany; Shanghai Frontiers Science Center of Genome Editing and Cell Therapy, Shanghai Key Laboratory of Regulatory Biology, Institute of Biomedical Sciences and School of Life Sciences, East China Normal University, 500 Dongchuan Road, Shanghai 200241, China; Shanghai Institute of Transplantation, 1630 Dongfang Road, Shanghai, 200127, China; Shanghai Engineering Research Center of Transplantation and Immunology, 1630 Dongfang Road, Shanghai, 200127, China; Institute for Experimental Virology, TWINCORE, Centre for Experimental and Clinical; Infection Research, a Joint Venture Between the Helmholtz Centre for Infection Research and the Hannover Medical School, Hannover, Germany; Institute for Pulmonary Rehabilitation Research and Department of Medicine, Pulmonary and Critical Care Medicine, Schoen Klinik Berchtesgadener Land, Schoenau am Koenigssee, Germany; Philipps-University Marburg, German Center for Lung Research (DZL) Universities of Giessen and Marburg Lung Center (UGMLC), Marburg, Germany; Paracelsus Medical University, Salzburg, Austria; Heinrich Heine University of Düsseldorf, Faculty of Medcine

## Abstract

**BACKGROUND:** Alpha-1 antitrypsin deficiency (AATD) caused by the homozygous SERPINA1 Z variant (PiZZ) is characterized by reduced circulating functional alpha-1 antitrypsin (AAT) and accumulation of mutant Z-AAT. YOLT-202 is a lipid nanoparticle-delivered adenine base-editing therapy designed to precisely correct the PiZ variant (E342K) while avoiding unintended bystander editing. We evaluated the safety and pharmacodynamic effects of a single dose of YOLT-202 in patients with AATD.

**METHODS:** In this ongoing, first-in-human, single-center, open-label, single-arm, dose-escalation study, adults with genetically confirmed PiZZ AATD and a serum total AAT concentration of less than 11 μM received a single intravenous dose of YOLT-202. In this interim analysis, one patient received 35 mg and three received 45 mg. The primary end point was safety and secondary end point was changes from baseline in serum total AAT concentrations.

**RESULTS:** Serum total AAT concentrations increased rapidly in all four patients, reaching or exceeding the putative protective threshold of 11 μM by week 3. At week 4, total AAT concentrations were 15.4 μM in the patient who received 35 mg and a mean of 19.0 μM in the 45-mg cohort, with increases sustained for up to 11 months. Circulating M-AAT concentrations increased, whereas Z-AAT concentrations decreased. At week 3, M-AAT accounted for 98.7% of total circulating AAT in the 35 mg patient and a mean of 93.4% in the 45-mg cohort. Adverse events occurred in two patients (50%), including infusion-related reactions in two and liver-enzyme elevations in two. No treatment-related adverse events of grade 3 or higher, serious adverse events, or deaths were reported.

**CONCLUSIONS:** In this small, ongoing study, a single administration of YOLT-202 was associated with sustained increases in circulating AAT concentrations and a shift from Z-AAT toward M-AAT. Preclinical findings and liver-biopsy results from one patient provided evidence of precise correction of the PiZ variant with negligible bystander editing. No serious adverse events or treatment-related adverse events of grade 3 or higher were observed. (ClinicalTrials.gov number, NCT07193615.)

---

Alpha-1 antitrypsin deficiency (AATD) is an inherited disorder caused by pathogenic variants in the *SERPINA1* gene, which encodes alpha-1 antitrypsin (AAT), a serine protease inhibitor primarily produced in the liver that protects tissues from neutrophil serine proteases (NSPs) such as neutrophil elastase (NE) and proteinase 3^1^. In AATD, impaired production or function of AAT results in unopposed protease activity in the lungs, leading to degradation of elastin and other extracellular matrix components in the alveoli, which manifests clinically as early-onset emphysema, bronchiectasis and chronic obstructive pulmonary disease; environmental exposures such as smoking and air pollution further accelerate this lung damage ^2^. Concurrently, polymerization of misfolded AAT in hepatocytes causes cellular injury, resulting in liver fibrosis, cirrhosis, and an increased risk of hepatocellular carcinoma^3^. The most common deficiency alleles are Z (E342K, corresponding to E366K in the full-length AAT precursor) and S (E264V, corresponding to E288V in the full-length precursor). The Z allele causing protein misfolding and polymerization. More than 95% of severe cases carry the homozygous PiZZ genotype, which occurs in approximately 1 in 5,000 European Caucasians^1,4^.

Current treatment strategies for AATD primarily aim to slow disease progression and manage organ-specific complications. Intravenous augmentation therapy with plasma-derived alpha-1 antitrypsin (AAT) is the only disease-specific pharmacologic treatment for AATD-related lung disease. In selected patients with severe AAT deficiency, augmentation therapy increases antiprotease capacity and slows the decline in lung density as measured by computed tomography^5,6^. Evidence regarding its effects on mortality and the frequency of exacerbations remains limited^7^. Augmentation therapy requires ongoing intravenous infusions and does not address the accumulation of misfolded Z-AAT in hepatocytes^8^. No disease-specific pharmacologic treatment has been approved for AATD-related liver disease. Management includes monitoring, supportive care, and liver transplantation for patients with end-stage liver disease^3,8^. Fazirsiran, an investigational RNA interference therapy, has been shown to reduce hepatic Z-AAT levels and improve measures of liver disease^9^. However, fazirsiran does not correct the deficiency of functional AAT that underlies AATD-related lung disease, a major cause of morbidity and mortality in adults with AATD^10^. Thus, direct correction of the pathogenic SERPINA1 Z allele could restore production of functional wild-type AAT (M-AAT) and reduce production of Z-AAT, potentially addressing both the pulmonary and hepatic manifestations of AATD.

Adenine base editing enables targeted A-to-G conversion without requiring DNA double-strand breaks and offers a potential approach to correcting the PiZ variant^11,12^. Some previously reported adenine base editors corrected the PiZ variant but also edited an adjacent adenine, introducing the D341G substitution (D365G in the full-length AAT precursor)^13–15^. Because the effects of D341G on therapeutic efficacy and safety remain uncertain, we developed an adenine base editor designed to minimize this bystander edit while maintaining efficient correction of the PiZ variant. We formulated the mRNA encoding the editor and a SERPINA1-targeting single-guide RNA (sgRNA) into lipid nanoparticles (LNPs) for in vivo delivery^16–18^. The resulting investigational therapy, YOLT-202, was designed to restore M-AAT production and reduce Z-AAT production after a single intravenous dose.

We report interim results from an ongoing dose-escalation study evaluating the safety and pharmacodynamic effects of a single intravenous dose of YOLT-202 in patients with AATD.

## METHODS

### PRECLINICAL STUDY

An adenine base editor was engineered and optimized specifically for correction of the pathogenic SERPINA1 PiZ variant to the wild-type sequence. Editor variants and sgRNAs targeting the PiZ locus were screened in PiZ HepG2 cells according to on-target correction and bystander editing. An sgRNA targeting the SERPINA1 PiZ locus (5′-AUCGACAAGAAAGGGACUGA-3′) was selected for use in YOLT-202. Editing activity was evaluated over a range of concentrations, and on-target correction, and bystander nucleotide substitutions were quantified by targeted next-generation sequencing. Potential genomic off-target sites were identified with the use of CALITAS, GUIDE-seq, and Digenome-seq. Candidate loci identified by these complementary approaches were subsequently evaluated by targeted deep sequencing in YOLT-202-treated PiZ HepG2 cells.

PiZ transgenic mice expressing human SERPINA1 E342K were used to assess the in vivo editing activity and pharmacodynamic effects of YOLT-202. Wild-type mice were used to assess safety.

### CLINICAL STUDY OVERSIGHT

The trial was sponsored by Ren Ji Hospital and funded by YolTech Therapeutics. The trial design and protocol were developed jointly by investigators at Ren Ji Hospital and Hannover Medical School and by medical experts from YolTech Therapeutics. Participants were recruited and screened at Hannover Medical School, and eligible participants received YOLT-202 at Ren Ji Hospital. The trial was conducted in accordance with the principles of the Declaration of Helsinki and the International Council for Harmonisation Good Clinical Practice guidelines, and the trial protocol was reviewed and approved by the Ethics Committee of Ren Ji Hospital, Shanghai Jiao Tong University School of Medicine. All participants provided written informed consent before enrollment. All participants provided written informed consent before enrollment. Data were collected by investigators at the participating academic institutions and by medical experts from YolTech Therapeutics. The authors had access to the study data, participated in the interpretation of the data and preparation of the manuscript, and vouch for the accuracy and completeness of the data and for the fidelity of the trial to the protocol.

### CLINICAL STUDY DESIGN AND ELIGIBILITY

Participants underwent baseline assessments during the screening period, including physical examination, vital-sign measurements, clinical laboratory testing, electrocardiography, pulmonary-function testing, and measurement of circulating AAT concentrations. Eligible participants received a single intravenous infusion of YOLT-202 at the assigned dose of 35 mg and 45 mg; the infusion was administered over a period of at least 2 hours. To reduce the risk of infusion-related reactions, participants received oral dexamethasone 8 to 24 hours before YOLT-202 administration and a glucocorticoid, a histamine H1-receptor antagonist, and a histamine H2-receptor antagonist approximately 1 hour before the infusion. Participants were monitored during and after the infusion for adverse events and clinically relevant changes in vital signs, laboratory values, and electrocardiographic findings. Participants underwent protocol-specified follow-up at Ren Ji Hospital through week 3 and at Hannover Medical School thereafter through week 52.

### CLINICAL ASSESSMENTS AND END POINTS

The primary end point was safety through week 52 after YOLT-202 administration, assessed on the basis of the incidence and severity of adverse events. This interim analysis included all safety data available through the data-cutoff date with follow-up ranged from 7 to 11 months. Safety assessments also included physical examinations, vital signs, electrocardiography, and clinical laboratory testing. Adverse events were graded according to the National Cancer Institute Common Terminology Criteria for Adverse Events, version 5.0.

### Statistical Analysis

Only descriptive analyses were planned, and no formal hypothesis testing was performed. Changes in circulating AAT protein levels from baseline are summarized descriptively as means, percentage changes from baseline, and ranges, as appropriate.

## RESULTS

### Preclinical study

Previous adenine base-editing strategies have enabled correction of the pathogenic SERPINA1 PiZ (E342K) variant but can also introduce unintended bystander substitutions (E341G)^13,14^. To improve editing precision, we further optimized our previously developed adenine base-editing platform, generating hpABE5-PiZM (Fig. 1B)^19^. In HepG2 cells engineered to harbor the SERPINA1 PiZ variant, on-target A-to-G editing was 11% at 1 ng and 92% at 32 ng of total RNA. Bystander editing remained below 1% at all doses tested (Fig. 1C).

**Figure 1.**
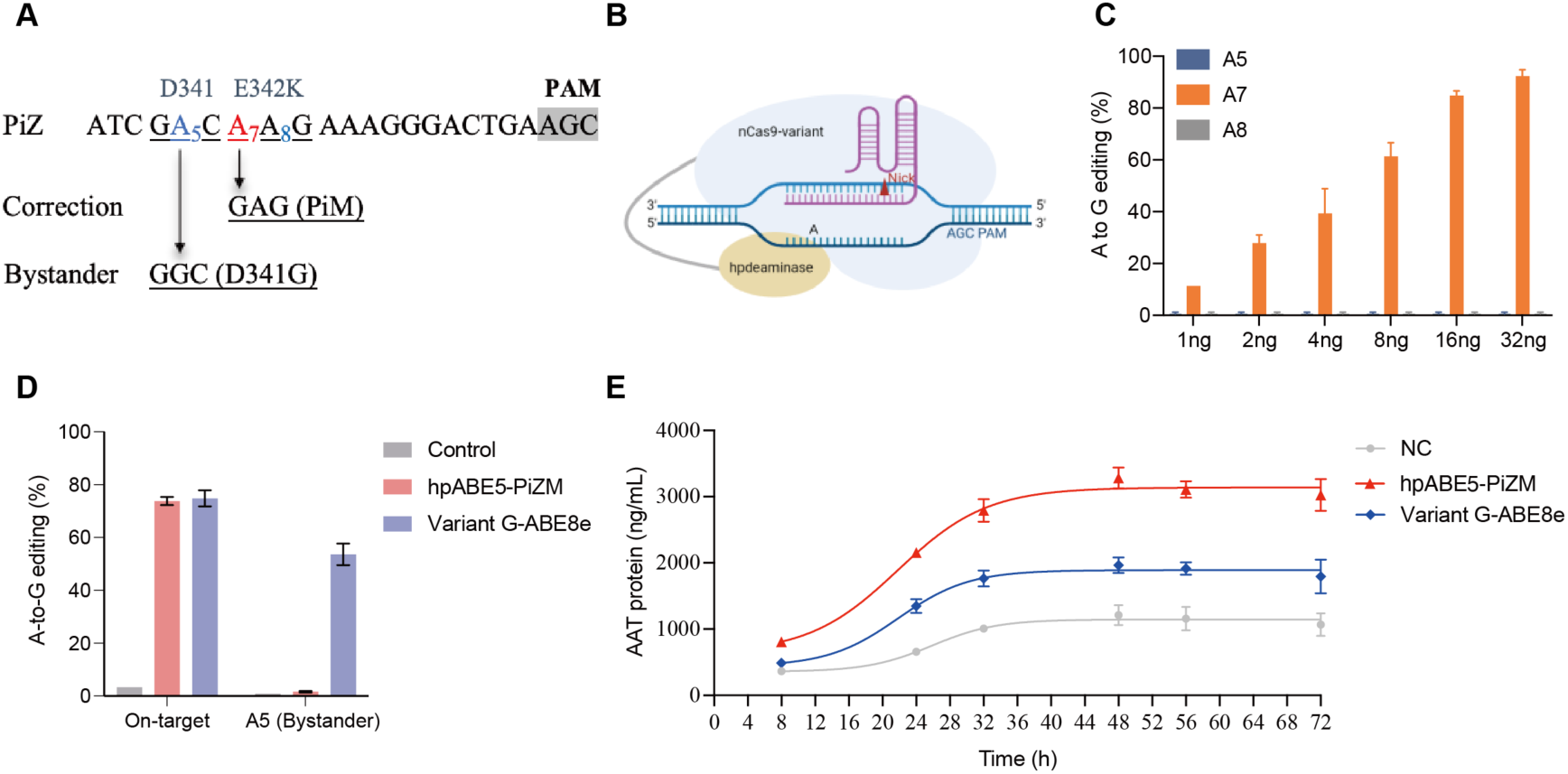
Precise correction of the SERPINA1 PiZ variant with hpABE5-PiZM. Panel A shows the SERPINA1 PiZ target sequence, including the pathogenic E342K mutation, the target adenine for correction, adjacent bystander adenines, and the protospacer-adjacent motif (PAM). Panel B shows a schematic of the hpABE5-PiZM base editor, consisting of an engineered adenine deaminase and a Cas9 nickase variant. Panel C shows dose-dependent A-to-G editing at the target adenine (A7) and adjacent bystander positions (A5 and A8) in HepG2 cells engineered to harbor the PiZ variant following treatment with increasing amounts of hpABE5-PiZM (1 to 32 ng). Panel D shows a comparison of on-target PiZ correction and A5 bystander editing in untreated control cells and cells treated with hpABE5-PiZM or Variant G-ABE8e. Figure E shows AAT protein concentrations in culture supernatants collected over 72 hours after medium replacement from untreated control cells and cells treated with hpABE5-PiZM or Variant G-ABE8e.

To determine whether bystander editing affects the functional restoration of AAT secretion following PiZ correction, we compared hpABE5-PiZM with Variant G-ABE8e, which was derived from Variant G disclosed in patent WO2024259364A2 by replacing its original deaminase with TadA-8e^11^. Both editors achieved E342K correction efficiencies exceeding 70% at the target adenine (A7) but differed markedly in editing at the adjacent bystander adenine (A5), which results in the unintended D341G substitution (Fig.1D). Despite comparable correction efficiencies, hpABE5-PiZM, which induced negligible bystander editing, resulted in substantially greater AAT secretion than Variant G-ABE8e. Approximately 40 hours after medium replacement, AAT secretion normalized to viable cell numbers increased to 2.60 times the untreated level with hpABE5-PiZM, as compared with 1.65 times with Variant G-ABE8e (Fig. 1E). These findings suggest that bystander editing may compromise the functional benefit of PiZ correction and underscore the importance of precise base editing for restoring AAT secretion.

We next evaluated potential off-target editing and structural variants following YOLT-202 treatment. Candidate sequence-dependent off-target sites were identified using three complementary approaches: CALITAS, GUIDE-seq, and Digenome-seq. A total of 7821 candidate sites were identified, most of which were nominated by computational prediction alone (Fig.S1A). Twenty-four sites were identified by all three approaches, and 124 (excluding the on-target locus) were identified by at least two approaches (Fig. S1B). These 124 sites were subsequently assessed by targeted deep sequencing in HepG2-PiZ cells treated with a saturating dose of YOLT-202. On-target editing at the PiZ locus reached 71.3%, whereas no treatment-associated A-to-G editing was detected at the evaluated candidate off-target sites (Fig. S2). We further assessed structural variants (SVs) using high-depth hybrid-capture next-generation sequencing (NGS) targeting the genomic region surrounding the SERPINA1 on-target site. No SV events with an allele frequency exceeding 0.1% were detected in YOLT-202-treated cells (Fig. S3).

In vivo editing and pharmacodynamic effects were evaluated in mice carrying the human *SERPINA1* PiZ transgene (Fig. S4A). Two weeks after a single intravenous dose of YOLT-202, mean hepatic PiZ correction was 22.6% at 0.03 mg per kilogram and ranged from 60.8% to 66.3% at doses of 0.05, 0.1, and 0.3 mg per kilogram. Bystander editing remained below 1% across all dose groups (Fig. S4B). PiZ correction was accompanied by increased serum AAT concentrations, from 24.1 μM in untreated controls to 81.2 and 95.0 μM at doses of 0.05 and 0.1 mg per kilogram, respectively (Fig. S4C). Liquid chromatography-mass spectrometry (LC-MS) analysis showed a corresponding shift from Z-AAT to M-AAT, with M-AAT accounting for 94.0% and 97.2% of total circulating AAT at the respective doses (Fig. S4D).

We further evaluated the tissue distribution of YOLT-202-mediated editing and its safety profile in PiZ mice. *SERPINA1* editing was predominantly hepatic, with editing efficiencies ranging from approximately 50% to 70% across the three dose groups and only low-level editing detected in extrahepatic tissues (Fig. S5). Transient elevations in serum alanine aminotransferase (ALT) and aspartate aminotransferase (AST) levels were observed after administration, peaking at 6 hours and subsequently declining toward baseline (Fig. S6B and S6B). Serum interleukin-6 (IL-6) levels also increased transiently, particularly in the highest-dose group, and declined toward baseline by day 1 (Fig. S7).

### Patients

A total of four patients were enrolled and received YOLT-202, including one in the 35-mg cohort and three in the 45-mg cohort. The first patient was enrolled on December 12, 2025, and the fourth on April 28, 2026. The patients ranged from 50 to 68 years of age, and three were men. Body weight ranged from 73.9 to 114.6 kg, and the body-mass index (BMI; the weight in kilograms divided by the square of the height in meters) ranged from 21.6 to 34.4. Two patients, both in the 45 mg cohort, had previously received augmentation therapy. Baseline serum AAT concentrations were below 11 μM in all four patients, with levels below the lower limit of quantification (3.68 μM) in three patients and a level of 8.05 μM in one patient. Liver stiffness, as measured by FibroScan, ranged from 5.8 to 7.1 kPa. Post-bronchodilator FEV1 ranged from 43 to 97% of the predicted value, and DLCO ranged from 26 to 74% of the predicted value. Additional baseline characteristics are shown in Table 1.

**Table 1.** Characteristics of the Participants at Baseline.

| Characteristic | 35 mg |  | 45 mg |  |
|---|---|---|---|---|
| Patient | S001 | S002 | S003 | S004 |
| Age range(yr) | 66-70 | 66-70 | 51-55 | 46-50 |
| Sex | Male | Male | Female | Male |
| *SERPINA1* genotype | PiZZ | PiZZ | PiZZ | PiZZ |
| Body Weight (kg) | 73.9 | 78.0 | 80 | 114.6 |
| Body Mass Index (Kg/m<sup>2</sup>) | 21.6 | 24.1 | 24.7 | 34.4 |
| History of Augmentation Therapy | No | Yes | Yes | No |
| Alpha 1 antitrypsin (AAT) (μM) | 3.68* | 8.05 | 3.68* | 3.68* |
| Median Fibroscan liver stiffness (kPa) | 5.8 | 7.1 | 5.9 | 6.5 |
| Actual FEV1 and %Pred | 2.63L; 70 | 1.51L;43 | 1.40L; 43 | 4.15L; 97 |
| Actual FEV1/FVC (%) and %Pred | 62.65; 82 | 41.86; 55 | 44.40; 55 | 76.38; 100 |
| Actual DLCO (mmol/min/kPa) and %Pred | 5.36; 66 | 2.06; 26 | 3.95; 54 | 7.24; 74 |

\* The lower limit of quantitation (LLOQ) of AAT at central lab using immunoturbidimetry is 3.68 μM. Results lower than LLOQ are displayed as 3.68 μM.

### Safety

All four patients received the intended dose of YOLT-202, and none withdrew from the trial. Adverse events occurred in two patients (50%). No serious adverse events or deaths occurred, and no treatment-related adverse events of grade 3 or higher were reported. The most common treatment-related adverse events were infusion-related reactions, which occurred in two patients (50%), and elevations in liver-enzyme levels, which occurred in two patients (50%) (Table 2). Mild elevations in body temperature (37.4°C and 37.3°C) were observed in two patients (50%) within 6 hours after infusion, with temperatures returning to normal within 2 hours after physical cooling measures (Fig. S8). Two patients (50%) had elevations in aminotransferase levels after infusion (the patient who received 35 mg and one of the three patients who received 45 mg). In both patients, aminotransferase levels had decreased to normal or near-normal levels by week 2 (Fig. S9A and S9B). Throughout follow-up, alkaline phosphatase levels were generally below the upper limit of the normal range (Fig. S9C), and total bilirubin levels remained normal in both patients (Fig. S9D).

**Table 2.** Summary of Safety and Adverse.

| Event | 35mg (n=1) | 45 mg (n=3) | Total (n=4) |
|---|---|---|---|
|  | n (%) | n (%) | n (%) |
| Any adverse event<sup>a</sup> | 1 (100) | 1 (33.3) | 2 (50) |
| Treatment-related adverse event (TRAE)<sup>b</sup> |  |  |  |
| Infusion-related reaction | 1 (100) | 1 (33.3) | 2 (50) |
| Liver enzyme elevation | 1 (100) | 1 (33.3) | 2 (50) |
| Any serious event | 0 | 0 | 0 |
| Any TRAE ≥ grade 3 in CTCAE severity | 0 | 0 | 0 |
| AE leading to discontinuation of YOLT-202 | 0 | 0 | 0 |
| AE leading to withdrawal from the trial | 0 | 0 | 0 |
| Death | 0 | 0 | 0 |

<sup>a</sup> The severity of adverse events was graded according to the National Cancer Institute Common Terminology Criteria for Adverse Events, version 5.0.
<sup>b</sup> The relatedness of adverse events to treatment was determined by the investigator.

### Efficacy

Total AAT concentrations increased in all four patients, exceeding the putative protective threshold of 11 μM by week 3 in the 35-mg patient and by week 2 in the 45-mg cohort. At week 4, the total AAT concentration was 15.4 μM in the patient who received 35 mg, and the mean concentration was 19.0 μM in the 45-mg cohort (Fig. 2). In the 45-mg cohort, mean total AAT concentrations approached the lower limit of the normal range (20 μM) from month 2 through month 7, with measurements available for all three patients at each visit. After the initial increase, total AAT concentrations were generally stable at elevated levels during follow-up in both dose cohorts. Measurements at month 11 were available for two patients, one in each cohort (Fig. 2).

**Figure 2.**
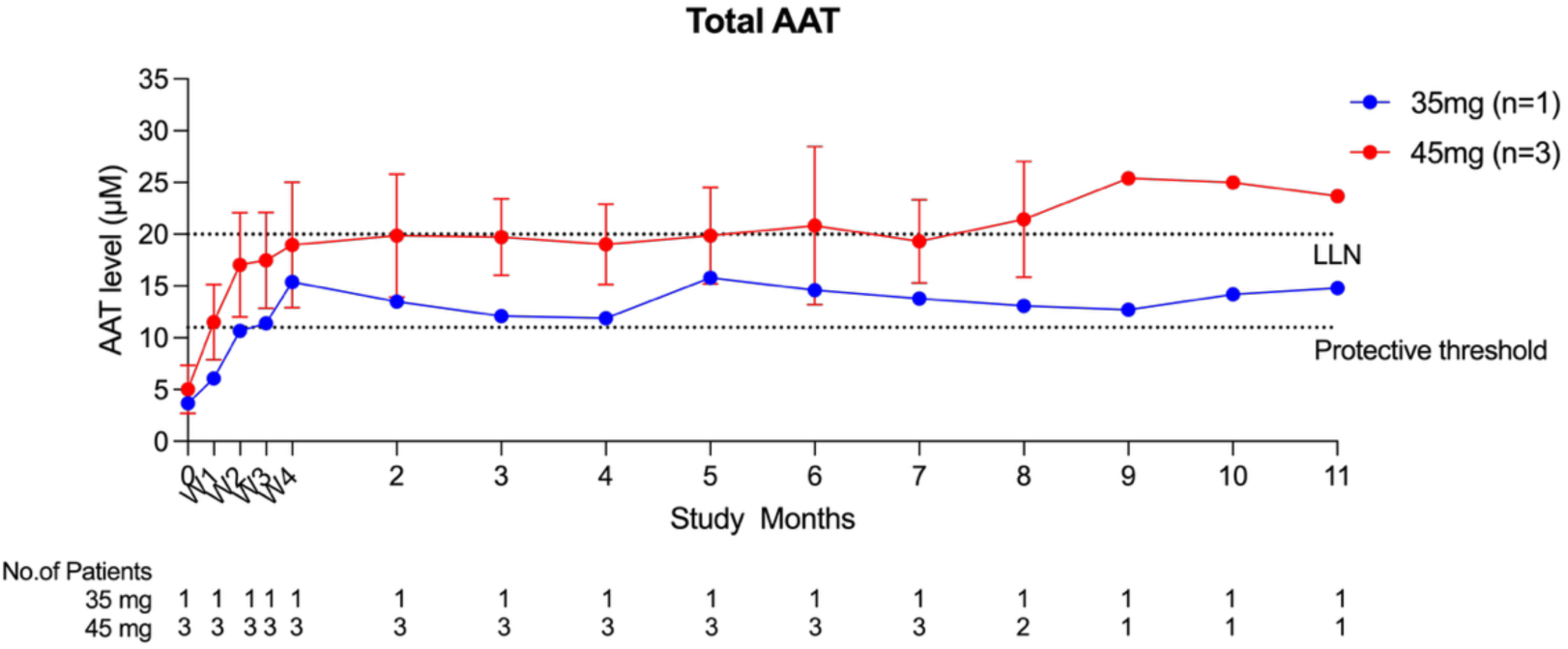
Serum total alpha-1 antitrypsin concentrations after a single dose of YOLT-202. Serum total AAT concentrations are shown over time for the patient who received 35 mg and as mean concentrations for the 45-mg cohort. Error bars indicate standard deviations. The numbers below the x-axis indicate the number of patients with available measurements at each visit. The horizontal dotted lines indicate the putative protective threshold (11 μM) and the lower limit of normal (LLN, 20 μM). W1-W4 denote weeks 1-4 after dosing.

Consistent with correction of the SERPINA1 PiZ variant, circulating wild-type M-AAT concentrations increased rapidly after YOLT-202 administration, whereas mutant Z-AAT concentrations decreased. At week 1, the M-AAT concentration was 6.6 μM in the 35-mg patient and averaged 12.8 μM in the 45-mg cohort, increasing to 15.0 μM and 21.9 μM, respectively, by week 3 (Fig. 3A). In parallel, Z-AAT concentrations decreased from baseline by 49.1% in the 35-mg patient and a mean of 69.4% in the 45-mg cohort at week 1, with corresponding reductions of 95.8% and 78.4% at week 3 (Fig. 3B). M-AAT accounted for 72.8% and 85.4% of total circulating AAT at week 1 in the 35-mg patient and the 45-mg cohort, respectively, increasing to 98.7% and 93.4% by week 3 (Fig. 3C). These changes resulted in a marked shift in circulating AAT composition from predominantly Z-AAT at baseline to predominantly M-AAT after treatment in both dose cohorts (Fig. 3D).

**Figure 3.**
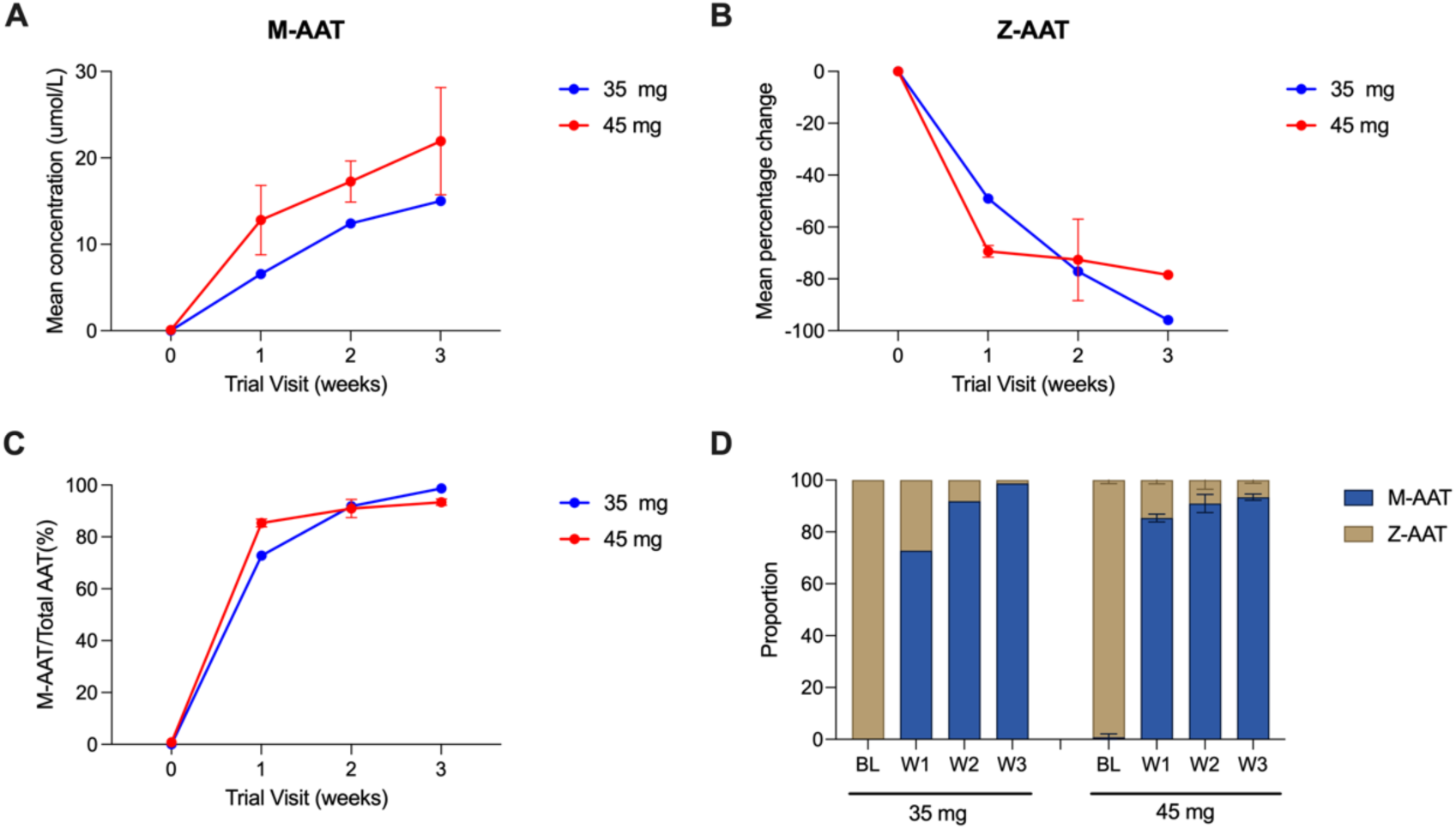
Serum M-AAT and Z-AAT after a single dose of YOLT-202. Panel A shows serum M-AAT concentrations from baseline through week 3. Panel B shows the percentage change from baseline in serum Z-AAT concentrations. Panel C shows M-AAT as a percentage of total serum AAT. Panel D shows the relative proportions of M-AAT and Z-AAT at baseline (BL) and at weeks 1, 2, and 3. Data are shown for the patient who received 35 mg and as means for the 45-mg cohort (n = 3 at weeks 1 and 2 and n = 2 at week 3).

## Discussion

In this interim analysis, a single intravenous dose of YOLT-202 was associated with rapid increases in total AAT and M-AAT concentrations. Total AAT concentrations exceeded the putative protective threshold of 11 μM in both dose cohorts, and circulating AAT shifted from predominantly Z-AAT at baseline to predominantly M-AAT after treatment^20^. These pharmacodynamic changes were consistent with the intended correction of the PiZ variant to the wild-type sequence and with the effects observed in preclinical cellular and animal models.

In this initial cohort of four patients, no serious adverse events or treatment-related adverse events of grade 3 or higher were observed. Infusion-related reactions and transient elevations in aminotransferase levels were observed, findings that have also been reported with other liver-directed lipid nanoparticle-based genome-editing therapies^21,22^. The infusion-related reactions were transient and resolved within 48 hours. The aminotransferase elevations were asymptomatic, were not accompanied by increases in bilirubin levels, and subsequently resolved. However, the small number of patients and the limited duration of follow-up preclude assessment of uncommon or delayed adverse events, and continued long-term follow up will be important.

YOLT-202 incorporates an adenine base editor optimized to correct the pathogenic E342K variant in SERPINA1. In preclinical studies, hpABE5-PiZM efficiently corrected E342K with negligible bystander editing in HepG2-PiZ cells and mouse models, despite the presence of multiple adenines within the editing window. Bystander editing at the adjacent Asp341 codon could affect the therapeutic benefit of PiZ correction. In a cell-based assay, the D341N substitution (D365N in the full-length AAT precursor) was associated with lower levels of secreted AAT monomer and lower neutrophil elastase inhibitory activity than wild-type AAT^23,24^. Kueppers et al. identified D341V in a heterozygous carrier with a serum AAT concentration within the range commonly observed in individuals with the PiMZ genotype and suggested, on the basis of structural modeling, that this substitution could disrupt the hydrophobic core of AAT^25^. Packer et al. reported comparable levels of D341G and wild-type AAT in the culture medium of transiently transfected HEK293T cells and similar elastase inhibitory activity, although the concentration-response curve for D341G AAT appeared shifted to the right^13^. In our liver-derived HepG2-PiZ model, AAT was expressed from the endogenous SERPINA1 locus under native promoter control.

Despite comparable rates of E342K correction, we observed lower AAT levels in the culture medium under editing conditions that introduced D341G than under conditions with negligible bystander editing. These findings support the development of precise editing strategies that correct E342K while avoiding bystander substitutions at Asp341.

In preclinical studies, no off-target editing was detected at the candidate sites evaluated in cultured cells. To assess editing specificity in vivo, liver biopsy was performed 10 weeks after YOLT-202 administration in the first treated patient (35 mg). Analysis of liver tissue showed correction of the pathogenic PiZ allele in 54% of alleles by Sanger sequencing and 57% by Illumina sequencing, with no detectable bystander or off-target editing^26^. These findings demonstrate the feasibility of precise, single-nucleotide correction of a disease-causing variant at its endogenous genomic locus in humans.

The observed pharmacodynamic effects suggest that direct correction of the PiZ variant may offer a therapeutic approach distinct from conventional AAT augmentation therapy, which increases circulating AAT without correcting the underlying genetic defect. Correction of the endogenous PiZ allele has the potential to address two components of AATD pathobiology: deficiency of functional circulating AAT, which contributes to pulmonary disease, and production of misfolded Z-AAT, which contributes to hepatic disease. The increases in M-AAT concentrations, together with the reductions in circulating Z-AAT, are consistent with these intended pharmacodynamic effects. However, whether these changes translate into clinical benefits for pulmonary or hepatic disease remains to be determined

This study has limitations. The number of patients was small, the study was open-label and uncontrolled, and the duration of follow-up was limited and differed among patients. Nevertheless, the increases in functional and total AAT and the shift from Z-AAT to M-AAT provide clinical evidence of the intended pharmacologic effect of YOLT-202. Continued follow-up will further characterize the durability of these effects and longer-term safety.

## Data Availability

All data produced in the present study are available upon reasonable request to the authors.

## Notes

### Competing Interest Statement

The authors have declared no competing interest.

### Clinical Trial

NCT07193615

### Author Declarations

Ethics committee of Ren Ji Hospital, Shanghai Jiao Tong University School of Medicine gave ethical approval for this work.

