## Supplementary Material for "Precise In Vivo Base-Editing Therapy for Alpha-1 Antitrypsin Deficiency"

### Supplementary Appendix

### INCLUSION AND EXCLUSION CRITERIA

Inclusion criteria

To be considered eligible to participate in this study, a subject must meet all the inclusion criteria listed below:

1. Male or female (inclusive) aged ≥ 18 and ≤ 70 years old at the time of signing informed consent.
2. Diagnosed with AATD and genetically confirmed as homozygous PiZZ mutation.
3. Blood total AAT level < 11 μM or equivalent protein in mg/dL.
4. Patients receiving augmentation therapy must be willing to discontinue it at least 6 weeks prior to signing the informed consent form (ICF) and throughout the study period, unless clinically indicated.

Exclusion Criteria

1. Body mass index (BMI) > 35 kg/$m^2$.
2. Patients who have undergone lung or liver transplantation, are on the waiting list for lung or liver transplantation, or have had lung volume reduction surgery (LVRS).
3. Clinical evidence of severe bronchiectasis, as judged by the investigator (e.g., excessive sputum production or recurrent infections requiring antibiotics [> 4 times per year]).
4. FEV1 ≤ 30% of predicted value after bronchodilator use at screening
5. Liver disease with any of the following:
    - Measured liver stiffness (FibroScan) ≥ 10 kilopascals (kPa).
    - Known history of hepatic cirrhosis or associated complications (e.g., varices, ascites, hepatic encephalopathy).
    - ≥ F2 hepatic fibrosis in patients with prior liver biopsy.
    - Alanine aminotransferase (ALT) or aspartate aminotransferase (AST) > 1.5 times the upper limit of normal (ULN).
    - Total bilirubin > ULN; or > 2 times the ULN if Gilbert's syndrome is documented.

- International normalized ratio (INR) ≥ 1.2 at screening. If deemed appropriate by the investigator and/or the prescribing physician, anticoagulants may be discontinued for a washout period or reversed with vitamin K. If necessary, a repeat INR < 1.2 is acceptable.
- Hepatitis B surface antigen (HBsAg) positive.
- Hepatitis C virus (HCV) antibody positive. If HCV antibody is positive, HCV RNA PCR must be negative.

6. Allergy to the drugs contained in lipid nanoparticles (LNP) or LNP-mRNA vaccines, or previous history of adverse reactions to LNP-based drugs;
7. Smoking more than 5 cigarettes per day or consuming the equivalent amount of nicotine or nicotine replacement products within the past 6 months before screening.
8. History of alcohol abuse within the past 6 months before screening (drinking more than 14 units of alcohol per week [1 unit ≈ 360 mL of beer or 45 mL of 40% alcohol or 150 mL of wine]); or positive alcohol breath test at the time of screening or admission.
9. Patients with poorly controlled hypertension by conventional treatments (systolic blood pressure [SBP] ≥ 180 mmHg and/or diastolic blood pressure [DBP] ≥ 110 mmHg).
10. Patients with poorly controlled diabetes (hemoglobin A1c ≥ 9%).
11. Presence of New York Heart Association (NYHA) class III-IV heart failure, left ventricular ejection fraction < 50%, or prolonged QTc interval (female > 470 ms, male > 450 ms) at the time of screening.
12. Myocardial infarction, unstable angina pectoris, percutaneous coronary intervention, coronary artery bypass grafting, severe deep vein thrombosis or pulmonary embolism within 3 months before screening; poorly controlled severe arrhythmias within the past 3 months before screening, such as poorly controlled recurrent and highly symptomatic ventricular tachycardia, rapid ventricular response atrial fibrillation, or supraventricular tachycardia; or planned cardiac surgery or cardiac revascularization during the main study period.
13. Cerebrovascular accidents within 6 months before screening;
14. Known or suspected systemic viral, parasitic, or fungal infections, or active infections requiring expected antibiotic therapy within 14 days after screening.

15. Human immunodeficiency virus (HIV) antibody positive at screening.
16. Anticoagulant therapy within 14 days before enrollment (such as warfarin, dabigatran, apixaban).
17. Patients with bleeding tendency or history of coagulation disorders (such as liver cirrhosis, malignant hematological diseases, antiphospholipid antibody syndrome).
18. Expected survival time less than 2 years.
19. History of malignant tumors within the past 5 years (excluding cured skin basal cell carcinoma, skin squamous cell carcinoma, cervical carcinoma in situ, low-grade prostate carcinoma in situ, and cured thyroid basal cell carcinoma).
20. History of drug abuse within the past 3 years.
21. Pregnant or lactating women.
22. Patients with other diseases in the blood system, digestive system, central nervous system, or endocrine system that the investigator believes will interfere with evaluation or limit participation in the trial.
23. Inability or unwillingness to cooperate or comply with pretreatment medication requirements or study procedures.
24. Any other condition deemed unsuitable for the clinical trial participation by the investigator.

### DRUG SUBSTANCES AND DRUG PRODUCT

The single-guide RNA (sgRNA) drug substance in the lipid nanoparticle (LNP) drug product comprised a 20-nucleotide spacer sequence corresponding to a protospacer DNA sequence within the SERPINA1 gene spanning the PiZ (E342K) variant and including the target adenosine nucleotide to be corrected by the drug product. The full-length sgRNA (5′-AUCGACAAGAAAGGGACUGAGUUUUAGAGCUAGAAAUAGCAAGUUAAAAUAA GGCUAGUCCGUUAUCAACUUGAAAAAGUGGCACCGAGUCGGUGCUUUU-3′) was chemically synthesized by GenScript and incorporated 2′-O-methyl and phosphorothioate modifications according to an optimized modification pattern, as previously described[1].

The messenger RNA (mRNA) drug substance in the lipid nanoparticle (LNP) drug product encoded hpABE5-PiZM, an adenine base editor developed for correction of the SERPINA1

PiZ variant. The mRNA comprised a 5′ cap, a 5′ untranslated region (UTR), the hpABE5-PiZM coding sequence, a 3′ UTR, and a 3′ polyadenylate tail. The coding sequence was codon-optimized to minimize uridine content, and uridine was replaced with N1-methylpseudouridine during in vitro transcription. Briefly, a plasmid DNA template containing the hpABE5-PiZM coding sequence and a 120-bp poly(A) sequence was linearized and transcribed with T7 RNA polymerase in the presence of ATP, CTP, GTP, N1-methylpseudouridine triphosphate, and a cap analogue.

The lipid nanoparticle (LNP) drug product was prepared using a modified microfluidic-mixing procedure based on a previously described method.37 The lipid phase consisted of an ionizable lipid, 1,2-distearoyl-sn-glycero-3-phosphocholine (DSPC), cholesterol, and DMG-PEG at a molar ratio of 50:10:38.5:1.5. The ionizable lipid used in this study is described in patent application WO2024152512A1 by YolTech Therapeutics. The hpABE5-PiZM mRNA and SERPINA1-targeting sgRNA were co-formulated in the same LNPs. The two RNA components were diluted in 50 mM citrate buffer (pH 4.0) at an mRNA:sgRNA mass ratio of 1:1 to generate the aqueous RNA phase. The total RNA dose was defined as the combined mass of hpABE5-PiZM mRNA and sgRNA. The aqueous RNA phase was mixed with the ethanolic lipid phase at an aqueous-to-ethanol volume ratio of 3:1 and a total flow rate of 12 ml per minute. LNPs used in nonhuman-primate and clinical studies were manufactured using the same formulation parameters with a scalable T-junction mixer–based system. The resulting LNPs underwent purification, buffer exchange, and concentration by tangential-flow filtration, followed by sterile filtration through 0.22-μm filters. Particle size, polydispersity index (PDI), and zeta potential were measured by dynamic light scattering with a Zetasizer Nano ZS instrument (Malvern Panalytical). RNA encapsulation efficiency was determined with the Quant-iT RiboGreen RNA assay (Life Technologies) by measuring RNA fluorescence before and after detergent-mediated disruption of the LNPs. Encapsulation efficiency was calculated as 100 x (total RNA−unencapsulated RNA)/total RNA. For each LNP batch used in the study, particle size, PDI, zeta potential, RNA encapsulation efficiency, total RNA concentration were recorded.

### PRE-CLINICAL ASSESSMENT OF OFF-TARGET EFFECTS OF YOLT-202

A comprehensive off-target profiling was performed to characterize the propensity of YOLT-202 to introduce editing events at non-target genomic loci. This assessment consisted of two sequential phases: identification of potential off-target sites and experimental validation.

In the identification phase, three complementary strategies were adopted for candidate locus identification. First, *in silico* prediction was carried out using the CALITAS tool against the human reference genome assembly GRCh38. Predicted off-target loci were evaluated for mismatches and bulge formations relative to the YOLT-202 sgRNA spacer sequence. Only loci with a combined total of fewer than five mismatches and bulges were retained in the curated list of putative off-target sites.

Second, Digenome-seq was applied for biochemical profiling of CRISPR-mediated off-target cleavage[2]. Purified human genomic DNA was incubated with YOLT-202 gRNA, hpABE5-PiZM and EndoV enzyme to induce targeted double-strand breaks (DSBs). The treated DNA was then subjected to whole-genome sequencing at 90× depth to map genome-wide cleavage sites. A blank control without gRNA supplementation was included to eliminate background signals derived from sample preparation artifacts and intrinsic sequencing noise.

Third, GUIDE-seq was utilized as a cell-based genome-wide approach to capture off-target editing events[3]. HepG2 cells were electroporated with YOLT-202 gRNA, Cas9 protein (the Cas9 variant component of hpABE5-PiZM), and synthetic double-stranded oligodeoxynucleotides (dsODNs). The dsODN fragments integrate specifically into CRISPR-induced DSBs, enabling unambiguous identification of both on-target and off-target loci. Sequencing libraries were constructed following standard protocols, and paired-end 150 bp sequencing was performed on Illumina MiSeq and HiSeq 2500 platforms.

In the validation phase, a modified liver-originated HepG2 cell line was used to assess on-target editing efficiency. A PiZ-mutant HepG2 cell line was established by introducing the SERPINA1 PiZ mutation into the endogenous wild-type SERPINA1 locus. A total of 124 putative off-target loci that were concurrently identified by no fewer than two independent screening methods were further validated in PiZ-mutant HepG2 cells. Cells were treated with a saturating dose of YOLT-202, and targeted deep sequencing was performed to quantify undesired off-target edits.

For structural variant (SV) analysis, hybrid-capture next-generation sequencing (NGS) was performed to interrogate an approximately 9-kb genomic region surrounding the SERPINA1 on-target site, enabling high-depth assessment of potential structural alterations associated with YOLT-202–mediated editing.

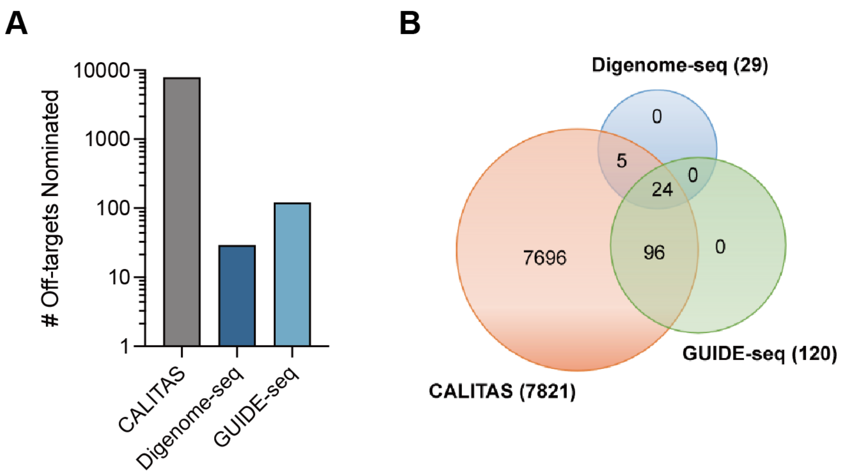


**Fig.S1 Determination of candidate off-target site list.**

Panel A shows the total candidate off-target sites quantified by each methodology at relevant nomination stringency thresholds. Panel B shows the overlap of candidate off-target sites identified by CALITAS, GUIDE-seq, and Digenome-seq.

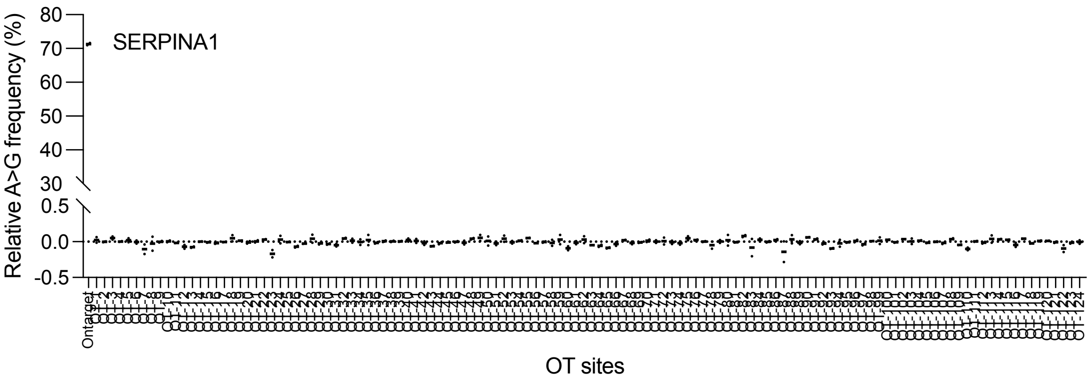


**Fig.S2 Verification of candidate off-target sites.**

A-to-G editing frequencies at the on-target SERPINA1 locus and 124 candidate off-target sites, as determined by targeted deep sequencing in untreated and YOLT-202–treated HepG2-PiZ cells. Data from two independent experiments are shown. At each candidate off-target site, the relative A-to-G editing frequency was calculated by subtracting the frequency in untreated cells from that in YOLT-202–treated cells.

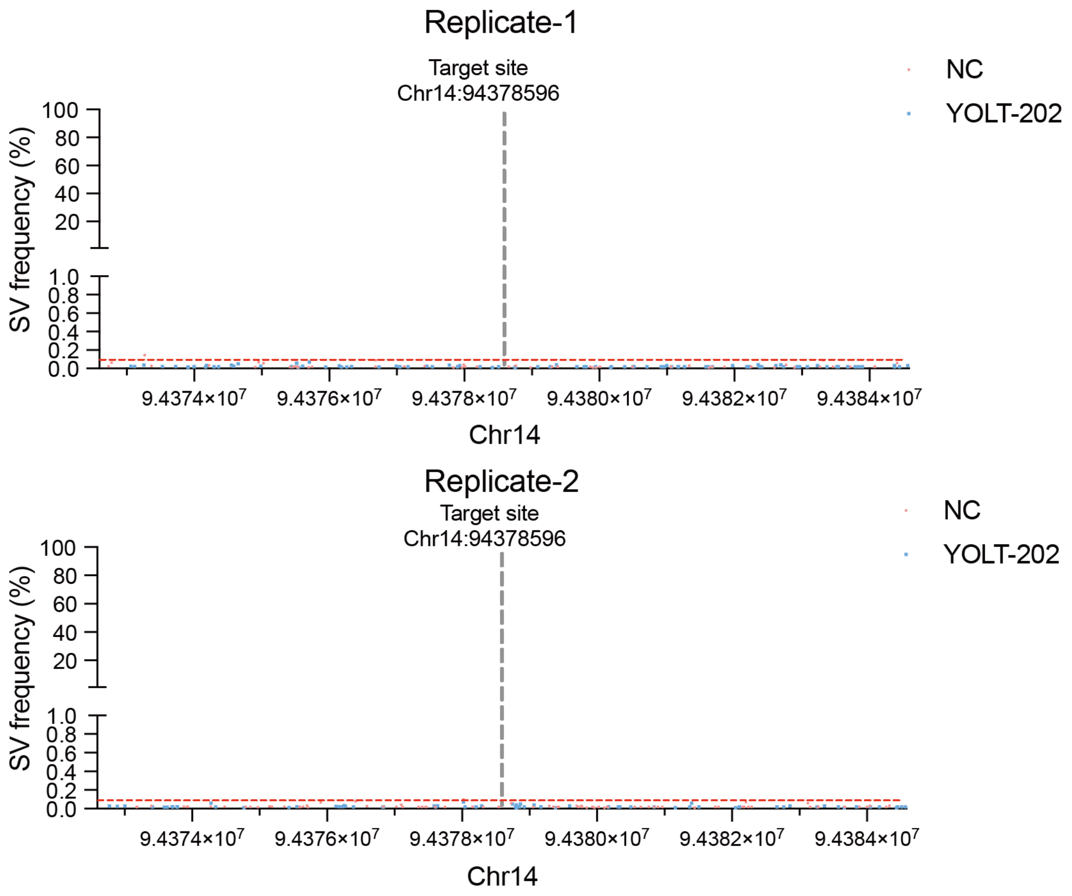


**Fig.S3 Structural variant analysis around the SERPINA1 PiZ target site after base editing.**

Structural variant (SV) frequencies across the genomic region surrounding the SERPINA1 target site on chromosome 14, as assessed by high-depth next-generation sequencing in untreated (NC, red) and YOLT-202–treated (blue) cells. Two independent replicates are shown. The vertical dashed line indicates the on-target editing site (chr14:94378596).

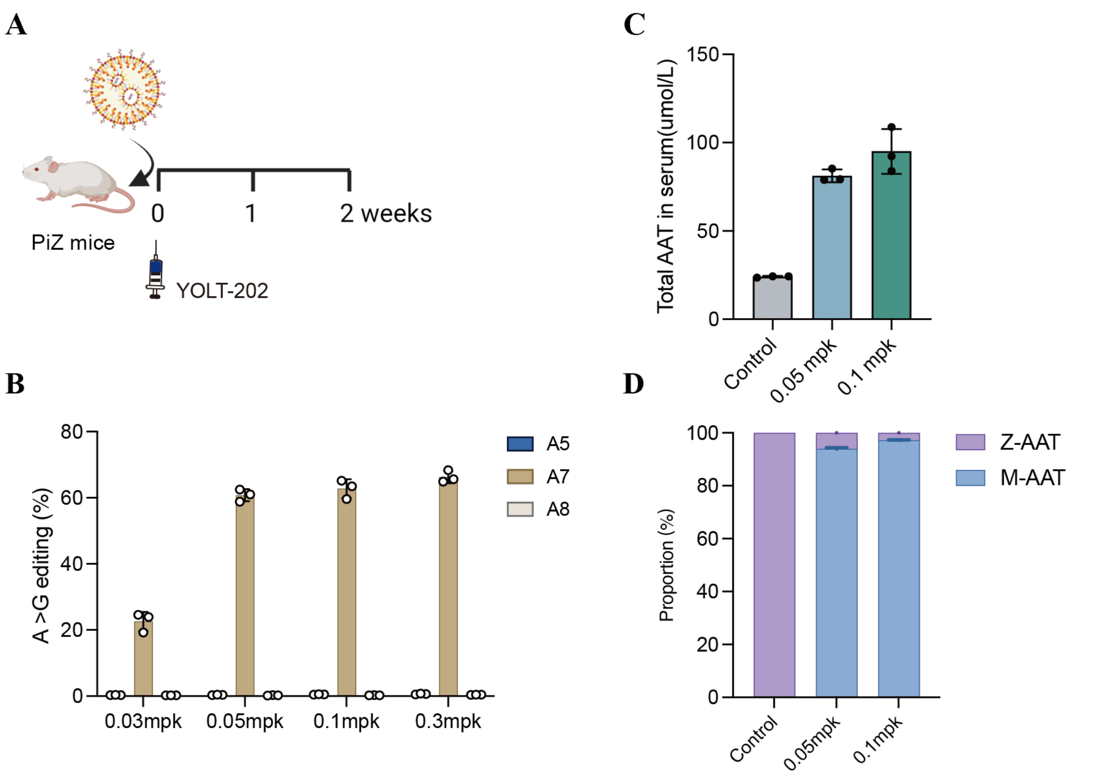


**Figure.S4 Preclinical animal study of YOLT-202.**

Panel A shows a schematic of the in vivo study design. PiZ mice received a single intravenous dose of YOLT-202 and were followed for 2 weeks. Panel B shows hepatic editing efficiencies at the target adenine (A7) and adjacent bystander adenines (A5 and A8) after administration of YOLT-202 at doses ranging from 0.03 to 0.3 mg per kilogram. Editing at A7 increased with dose and approached saturation at 0.1 mg per kilogram, whereas bystander editing at A5 and A8 remained minimal across all doses. Panel C shows serum total alpha-1 antitrypsin (AAT) concentrations before and after treatment with YOLT-202 at doses of 0.05 and 0.1 mg per kilogram. Panel D shows the relative proportions of circulating Z-AAT and M-AAT before and after treatment, as determined by liquid chromatography–mass spectrometry (LC–MS).

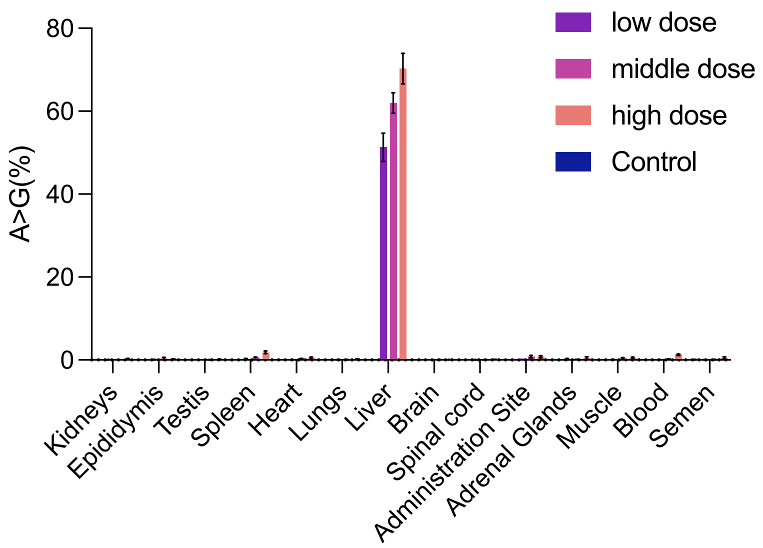


**Figure S5 Tissue distribution of A-to-G editing after YOLT-202 administration in mice.**

A-to-G editing at the target site was measured in the indicated tissues from mice in the low-, middle-, and high-dose groups and in control mice. Data are presented as means ±SD.

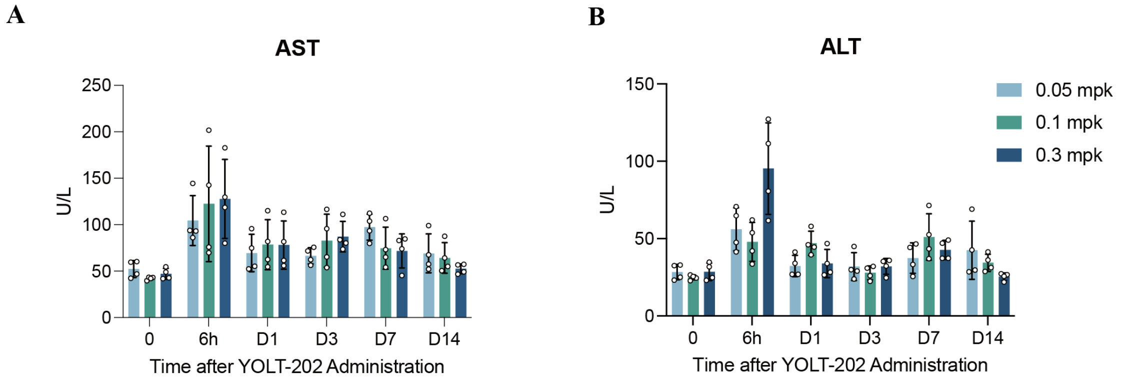


**Figure S6. Serum aminotransferase levels after YOLT-202 administration in mice.**

Panel A shows serum aspartate aminotransferase (AST) levels, and Panel B shows serum alanine aminotransferase (ALT) levels in mice that received a single intravenous dose of YOLT-202 at 0.05, 0.1, or 0.3 mg per kilogram. Serum levels were measured at baseline and at the indicated time points after dosing. Data are shown as means ± SD. Each dot represents an individual mouse.

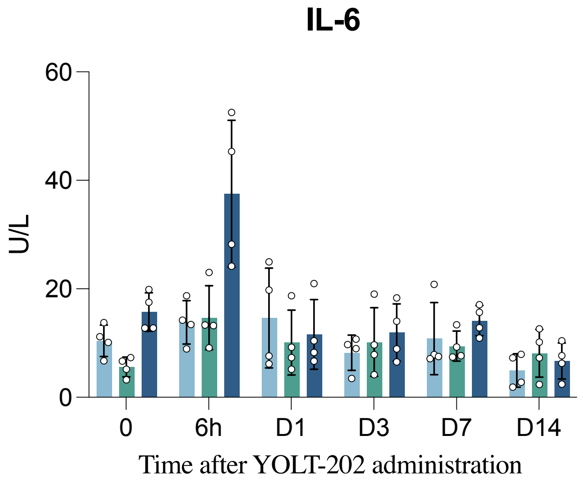


**Figure S7. Serum IL-6 Levels after YOLT-202 administration in mice.**

Serum interleukin-6 (IL-6) levels were measured in mice receiving a single intravenous dose of YOLT-202 at 0.05, 0.1, or 0.3 mg per kilogram. Levels were assessed at baseline and at 6 hours and 1, 3, 7, and 14 days after administration. Data are presented as means ±SD; dots represent individual animals.

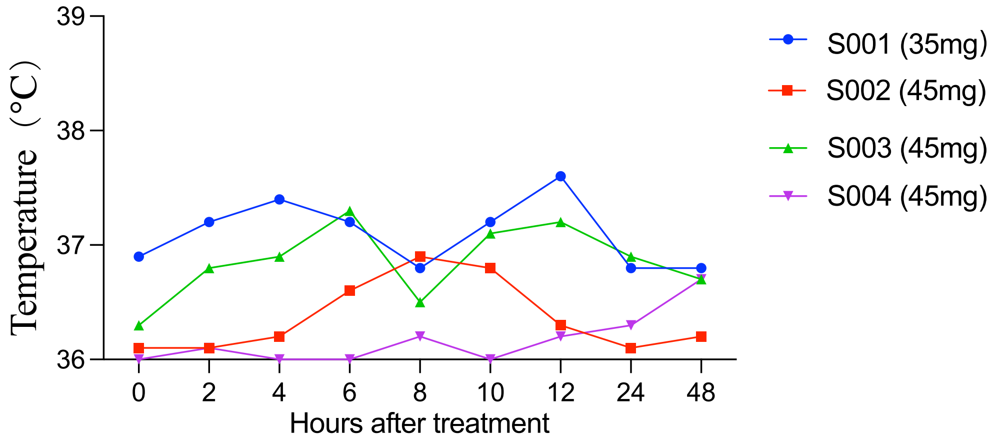


**Fig.S8 Body temperature during the first 48 hours after YOLT-202 administration.** Body temperature is shown from baseline through 48 hours after dosing. Each line represents one patient. One patient received 35 mg of YOLT-202, and three received 45 mg.

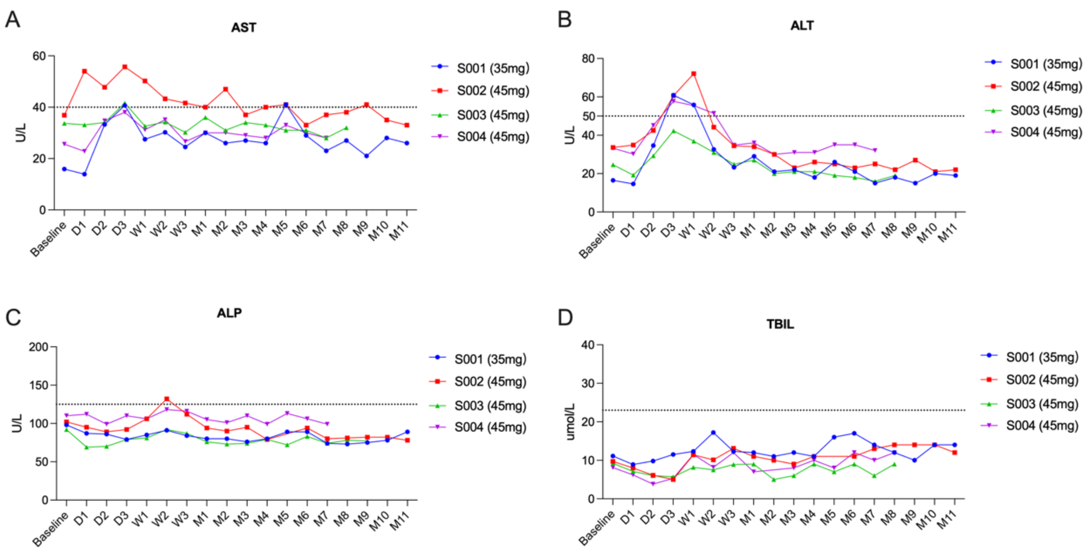


**Figure S9. Liver biochemical measures after YOLT-202 treatment.**

Shown are individual values for aspartate aminotransferase (AST) (Panel A), alanine aminotransferase (ALT) (Panel B), alkaline phosphatase (ALP) (Panel C), and total bilirubin (TBIL) (Panel D) from baseline through follow-up after a single intravenous dose of YOLT-202. Each line represents an individual participant. Dotted horizontal lines indicate the upper limit of the normal range.
